# Coronavirus and Post-COVID-19 Syndrome: A Systematic Review

**DOI:** 10.1101/2023.09.24.23296022

**Authors:** Sarvinoz Albalushi, Azmaeen Zarif, Suheyla Karaduman, Alesia Talpeka, Khoa Tran

## Abstract

Coronavirus infectious Disease 2019 (COVID-19) was first reported in Wuhan, China, and with its rapidly mutating variants, it soon became a global concern. In response to the pandemic, intensive research and development efforts led to the development of six vaccines approved by the World Health Organization (WHO). Coronavirus is divided into four genera: alpha, beta, gamma and delta. Its unstable ssRNA resulted in multiple strains in a short period, which acted as a selection pressure for transmissibility. Sequelae of COVID-19 infection include multiple syndromes which have been reported at high incidence globally. Using the Cochrane guidelines and the Preferred Reporting Items for Systematic Reviews and Meta-Analyses (PRISMA), we present a systematic review of the most common syndromes reported. A total of 12 eligible studies were included in this review. Syndromes reported in the literature include immune thrombocytopenic purpura (ITP), viral encephalomyelitis, hemophagocytic lymphohistiocytosis, thrombotic thrombocytopenic purpura (TTP), Guillain-Barrè syndrome (GBS) and postural orthostatic tachycardia syndrome (POTS). We cover the hypothesized pathophysiology, presenting symptoms and treatment for each respective syndrome. We aim to discuss coronavirus and its variants to provide a foundation on which to examine the syndromes manifested after COVID-19 infection (post-COVID-19 syndrome).

## Introduction

### Background

The first cases of Coronavirus infectious Disease 2019 (COVID-19), an infectious disease caused by the SARS-CoV-2 virus, were reported in Wuhan, China in late December 2019 (Li et al., 2020; Roy, Roy & Paul, 2020). The first four cases were reported as respiratory illnesses and initially referred to as ‘pneumonia of unknown etiology’, which later emerged as the global COVID-19 pandemic (Tan et al., 2020). Since its emergence, several variants have emerged. Despite strict preventive measures (sanitizing probable fomites, wearing face masks, regular hand washing, promoting personal hygiene, and using alcohol-based hand sanitizers) recommended by the Centers for Disease Prevention and Cure (CDC) and World Health Organisation (WHO), the number of affected individuals till date has reached 695 million where 6.9 million individuals died and 667 million recovered (Worldometer. (n.d.)).

In response to the pandemic, groundbreaking efforts were made by scientists in collaboration with pharmaceutical companies to develop six COVID-19 vaccines approved by the WHO’s Strategic Advisory Group of Experts on Immunization (SAGE). These vaccines included BNT162b2 mRNA (Pfizer), mRNA-1273 (Moderna), and Janssen Ad26.CoV2.S (Johnson & Johnson), AZD1222 (Oxford/AstraZeneca), CoronaVac (Sinovac Biotech), and BBIBP-CorV (Sinopharm). According to the WHO, by March 2022, 90% of the global population had antibodies against COVID-19 disease either through infection or through vaccination (WHO, 2022a). Alongside the development and mass administration of vaccines, several mutants of SARS-CoV-2 have been identified across the globe, which resulted in subsequent infection cycles and an exponential expansion in the geographical range of the disease.

Coronaviruses are a group of single-stranded RNA (ssRNA) viruses that fall under the family Coronaviridae, subfamily Coronavirinae, and order Nidovirales. These viruses transmit infection from human to human and also infect birds (Yousefi & Eslami, 2022). These viruses were documented almost 50 years ago and over these five decades, different viruses from this family have been identified in different areas of the globe. The viruses can be divided into four different genera: *Alpha coronavirus* (also known as human coronaviruses, HCoVs), *Beta coronavirus* (this category includes Middle Eastern Respiratory syndrome-related coronavirus, MERS-CoV, and the widely-known SARS-CoV), *Gamma coronavirus*, and *Delta coronavirus*. The viruses from the last two genera mostly infect birds (Nakagawa, Lokugamage, & Makino, 2016). Based on the similarity of a sequence of nucleic acid, the newly identified Severe acute respiratory syndrome, coronavirus 2 (SARS-CoV-2) can be categorized as beta coronavirus (Xiao et al., 2020).

### Variants of Coronavirus

SARS-CoV-2 is an RNA virus with an unstable genome. This poses a constant threat of developing mutant strains that may become dominant through natural selection, often with higher transmissibility than previous strains.

Throughout the pandemic, several variants of SARS-CoV-2 have emerged. These mutations can be caused by environmental mutagens such as metal ions and UV radiation, as well as endogenous components of the virus (Sanjuán & Domingo-Calap, 2016). Typically, RNA viruses evolve gradually over time (Morley & Turner, 2017). During the pandemic, the WHO collaborated with national and international researchers to monitor the changes in the patterns of transmission of SARS-CoV-2, its clinical presentation, severity, and impact on public health measures. The WHO established close monitoring networks to identify potential Variants of Interest (VOIs) and Variants of Concern (VOCs) and to assess their risk to public health (WHO, 2022b). To date, four VOCs have been identified: Alpha, Beta, Gamma, and Delta.

The Beta variant (B.1.351) was first identified in South Africa in May 2020. This variant has been found to affect younger people more severely than previous strains. Since its initial detection, it has been identified in 80 countries and was thought to be the main cause of the third wave of the pandemic in South Africa. Of particular concern is the E484K mutation found in this variant, which allows it to evade the immune system more easily (CDC, 2020).

The U.K. was the first location where the Alpha variant (B.1.1.7) was identified., and it was the prime cause of the third wave in the U.K. This strain is not only highly transmissible but it is extremely lethal. It is 30-70% more transmissible and lethal than the original strain (Raheem et al., 2021). Note that these strains do not result from a single mutation but rather are the result of, to date, 149 mutations that have been identified in more than a hundred sequenced strains since the beginning of the pandemic till May 2020 (Awadasseid et al., 2021). These mutations usually change the spike proteins of the virus. These spike proteins are used by the virus to enter the cells in humans using the angiotensin-converting enzyme 2 (ACE2) receptor (Casalino et al., 2020). The first notable strain of the novel coronavirus, which became a predomination globally, was the D614G variant, also known as G614. This variant was the result of a missense mutation (23403A>G) of the S1 subunit (spike protein component). This mutation changed the original strain (D614) that was identified in Wuhan. It was originally spotted in Europe and, within a month, dominated the globe. The G614 variant had higher transmissibility than the original Wuhan variant or any other variant (Korber et al., 2020). In addition to having a higher transmissibility, it was also found to enhance the risk of death among individuals (Challen et al., 2021; Davies et al., 2020).

The Gamma variant (P.1) was first identified in Brazil in November 2020. It is twice as transmissible as the previous strain and was the primary cause of the second deadly wave of the coronavirus. Studies have shown that existing vaccines provide only 54-79% protection against this variant (WHO, 2022c).

The Delta variant (B.1.617.2) was first reported by the WHO on June 21, 2021. It is highly contagious and, to date, the fastest-spreading strain. This variant targets vulnerable populations, especially in areas with low COVID-19 vaccination rates. It was first identified in India, where it was 60% more transmissible than other strains and posed a higher risk of reinfection (Cherian et al., 2021). The Delta variant drove the second deadly wave of Coronavirus in India in the summer of 2021, affecting vulnerable individuals with pre-existing conditions (Mallapaty et al., 2021). It spread to 92 countries, accounting for 20% of new cases in the US and 60% of new infections in the UK (Mishra et al., 2021). A mutation in the spike protein of the Delta virus has led to a new variant known as Delta plus, which has been found in several countries, including Nepal, Portugal, the US, the UK, and Russia (Kupferschmidt & Wadman, 2021).

The Omicron variant (B.1.1.529) was first reported by WHO on November 24, 2021, and just two days later it was declared a variant of concern due to its increased detection in the South African region and the identification of several mutations in the spike protein, particularly in the immunogenic region (WHO, 2021). This variant has nearly 32 mutations in the spike protein and 10 mutations in the receptor-binding domain. It was first identified in the U.S. and has since spread rapidly throughout the world. The mutations in the spike protein have made it easier for the virus to infect human cells and evade the immune response, making it more transmissible than previous strains. The most notable mutations are N501Y, D614G, and E484K, which allow the virus to escape the protection offered by currently available vaccines. These structural changes also distort the antibody binding sites, further compromising the immune response to the infection (Kamble et al., 2022).

### Composition and Genome Structure of Coronavirus

The family Coronaviridae encompasses the Coronavirus, which is further divided into two subfamilies: Coronavirinae and Torovirinae. ssRNA and +ssRNA are about 26–32 kb in genome size. Coronaviruses are typically classified into four genera based on their genomic structure and host range. The four genera of Coronaviridae are Alpha, Beta, Gamma, and Delta Coronavirus. The Alpha and Beta genera primarily infect mammals, while the Gamma and Delta genera primarily infect birds. Bats are the natural reservoir of Coronaviruses, and they are mainly responsible for the spread of novel coronavirus (Li et al., 2019). Coronavirus has at least 27 proteins including four structural proteins, 15 non-structural proteins, and 8 auxiliary proteins. Its envelope includes four spike-shaped proteins including envelope proteins (E), glycoproteins (S), nucleocapsid (N), and membrane proteins (M). These proteins play a vital role in binding to the host cell and facilitating entry of the virus. Sixteen non-structural proteins (nsp1–16) constitute a viral transcriptase-replicase complex that is encoded by two polypeptides (pp1a and pp1ab). These non-structural proteins are primarily involved in the formation of double-membraned vesicles that are derived from the rough endoplasmic reticulum, which plays a crucial role in the replication and transcription of the virus, serving as the primary site for these processes.

To reduce the number of mutations in the RNA genome, Coronavirus also encodes exoribonuclease (EXoN) which is produced by the nsp14 protein. The spike protein is a key factor in the virus’s ability to enter host cells, with the receptor-binding domain (RBD) located in the S1 subunit responsible for binding to the host cell receptor and the S2 subunit facilitating fusion of the viral and host cell membranes (Chen, Liu & Guo, 2020).

Spike glycoprotein (S), a structural protein that is located on the outer envelope of the virus, attaches to the host-receptor angiotensin[converting enzyme 2 (ACE2). The S protein of Coronavirus contains carboxyl (C)-terminal S2 subunit and amino (N)-terminal S1 subunit with a length ranging from 1104 to 1273 amino acids. The S1 subunit is the receptor-binding domain (RBD) which has an external subdomain and a core subdomain. These two subunits span around 200 residues. The core subdomain of RBD is primarily responsible for the formation of S trimer particles. The two exposed loops on the surface of the external subdomain are responsible for binding with ACE2.

Infection with COVID-19 has resulted in many different post-infective syndromes being reported, termed post-COVID syndrome. We attempted to systematically synthesize all the evidence available on sequelae syndromes post-COVID-19 infection.

## Method

The guidelines of the Cochrane Handbook for a systematic review and the Preferred Reporting Items for Systematic Reviews and Meta-Analyses (PRISMA) (Vrabel, 2015) statement were followed to structure this systemic review (Higgins et al. 2019).

### Search Strategy

A search strategy was established based on key terms for ‘Syndrome’ and ‘COVID-19’. Moreover, MeSH terms were reviewed to add a term to the search strategy.

PubMed was searched using the search strategy given in Table 1 to ensure all the studies conducted were captured. The search was limited to the last 4 years from 2019 (the emergence of COVID-19) to 2022.

**Table 1.** Search strategy for the review.

| Step | Term | Search term |
| --- | --- | --- |
| Step 1 | Syndrome | ‘Syndromes’ OR ‘Symptom Cluster’ OR ‘Symptom Clusters’ |
| Step 2 | COVID-19 | (‘COVID’ OR ‘COVID-19’ OR ‘nCoV’ OR ‘coronavirus’ OR ‘Middle East respiratory syndrome’ OR ‘MERS’ OR ‘Severe Acute Respiratory Syndrome’ OR ‘SARS’) |

### Screening Process

Searches were downloaded in the form of an Excel sheet. Duplicates were removed from the downloaded searches. Titles and abstracts were screened based on inclusion and exclusion criteria (see below). Then, full-length articles were screened using the same criteria. The PRISMA flowchart summarizes the screening and selection process of the studies (see Supplementary Materials, Figure S1).

### Determining the eligibility of the studies

Inclusion and exclusion criteria to determine the eligibility of screened studies for inclusion in the review are detailed below.

Inclusion Criteria:

- All quantitative studies, case reports, and case series reports on any syndrome that occurred after COVID-19 infection
- Both males and females from any age group were included
- Participants must have been diagnosed with COVID-19 infection
- Non-English studies were included if a full-text article was available in English

Exclusion Criteria:

- Qualitative studies, literature reviews, editorials, and policy documents were excluded
- Studies conducted on patients with any other infection than COVID-19
- Studies not having the outcome of interest (i.e., post-COVID-19 infection syndrome)
- No full text of the study was available

Inclusion and exclusion criteria based on intervention, comparator, population, and outcome of review question are detailed in the Supplementary Materials (Table S1).

### Risk of Bias

The risk of bias in studies was assessed using the Joanna Briggs Inventory (JBI) for case reports (see Supplementary Materials, Table S2) as case reports were the predominant type of manuscript expected for the given review question. JBI for case reports assesses the risk of bias in included studies using eight questions. These questions assess if: the patient’s demographics are clearly defined; there is an adequate description of the patient’s history; the current condition is described comprehensively; results of diagnostic tests and assessment methods are mentioned clearly; there is a clear description of treatment or intervention procedures; there is a comprehensive assessment of post-intervention clinical condition; and unanticipated or adverse events are adequately reported. Each question is answered in terms of yes, no, unclear, and not applicable (Munn et al., 2020).

### Data Extraction

After screening, manuscript data were extracted. Extracted data (see Table 2) included: author, year of publication, sample characteristics (including sample size, gender percentage of sample, mean age of the sample, or any other sample-specific characteristics), study design, study characteristics (outcome of the study), and results.

**Table 2:**
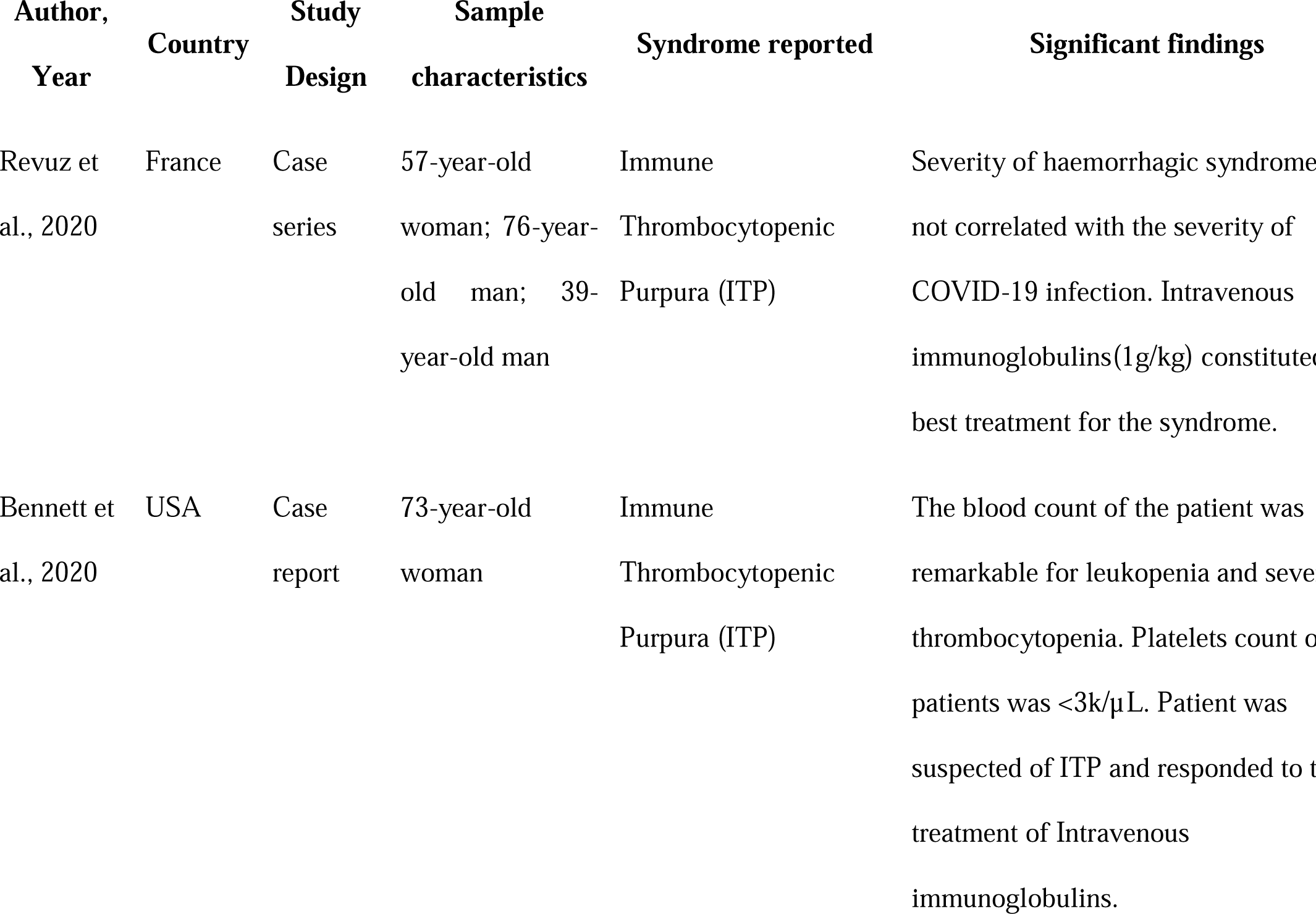
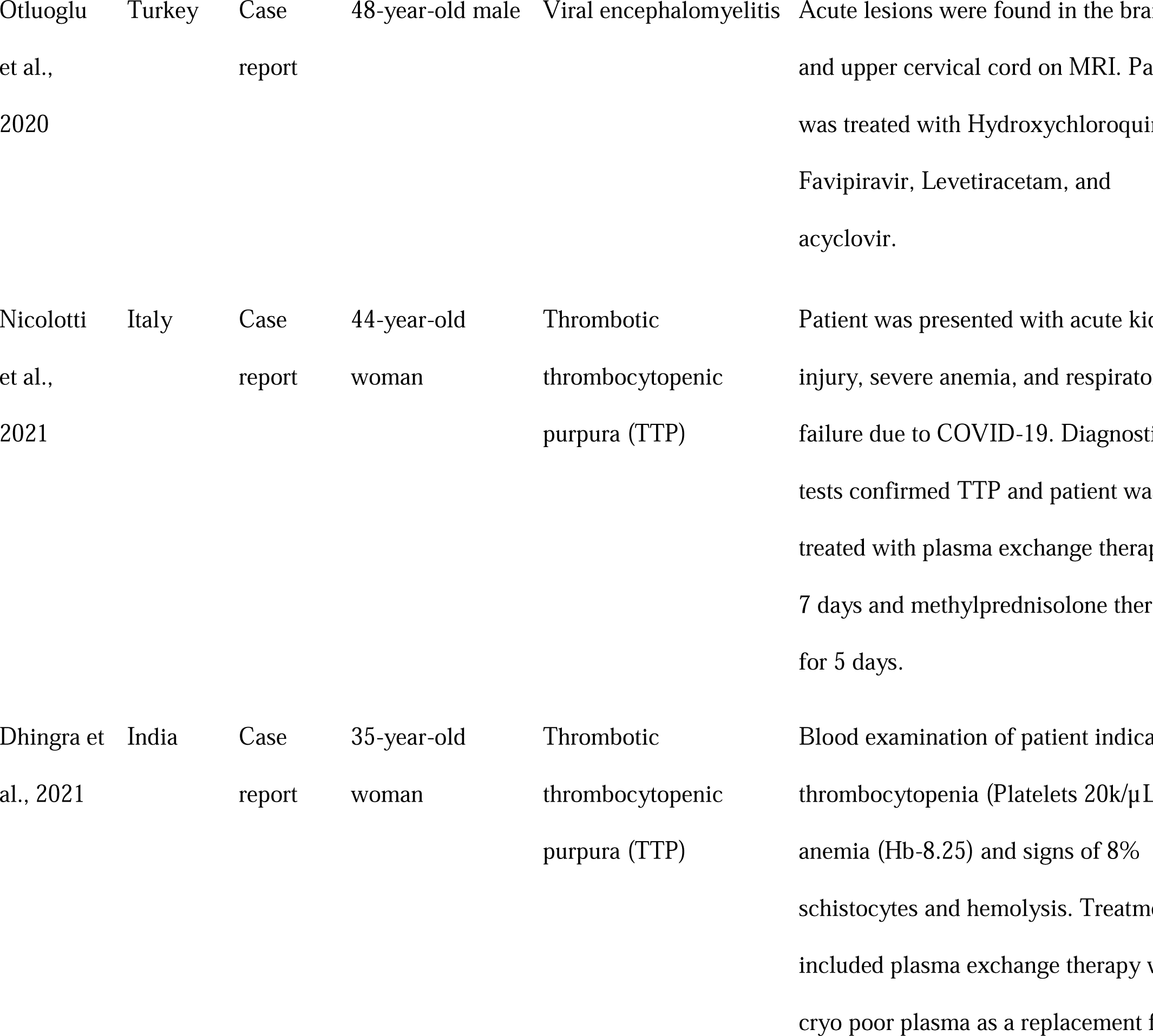
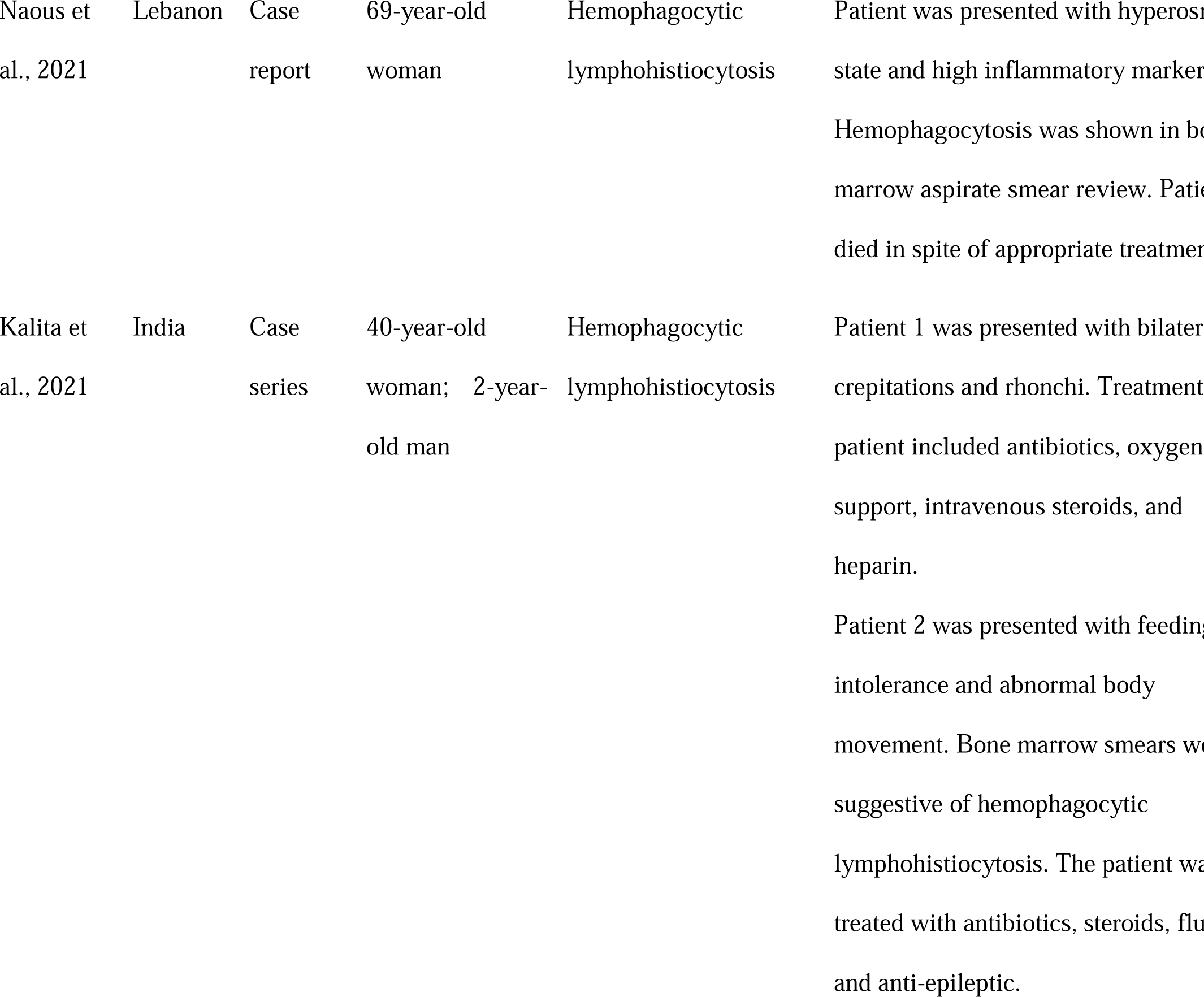
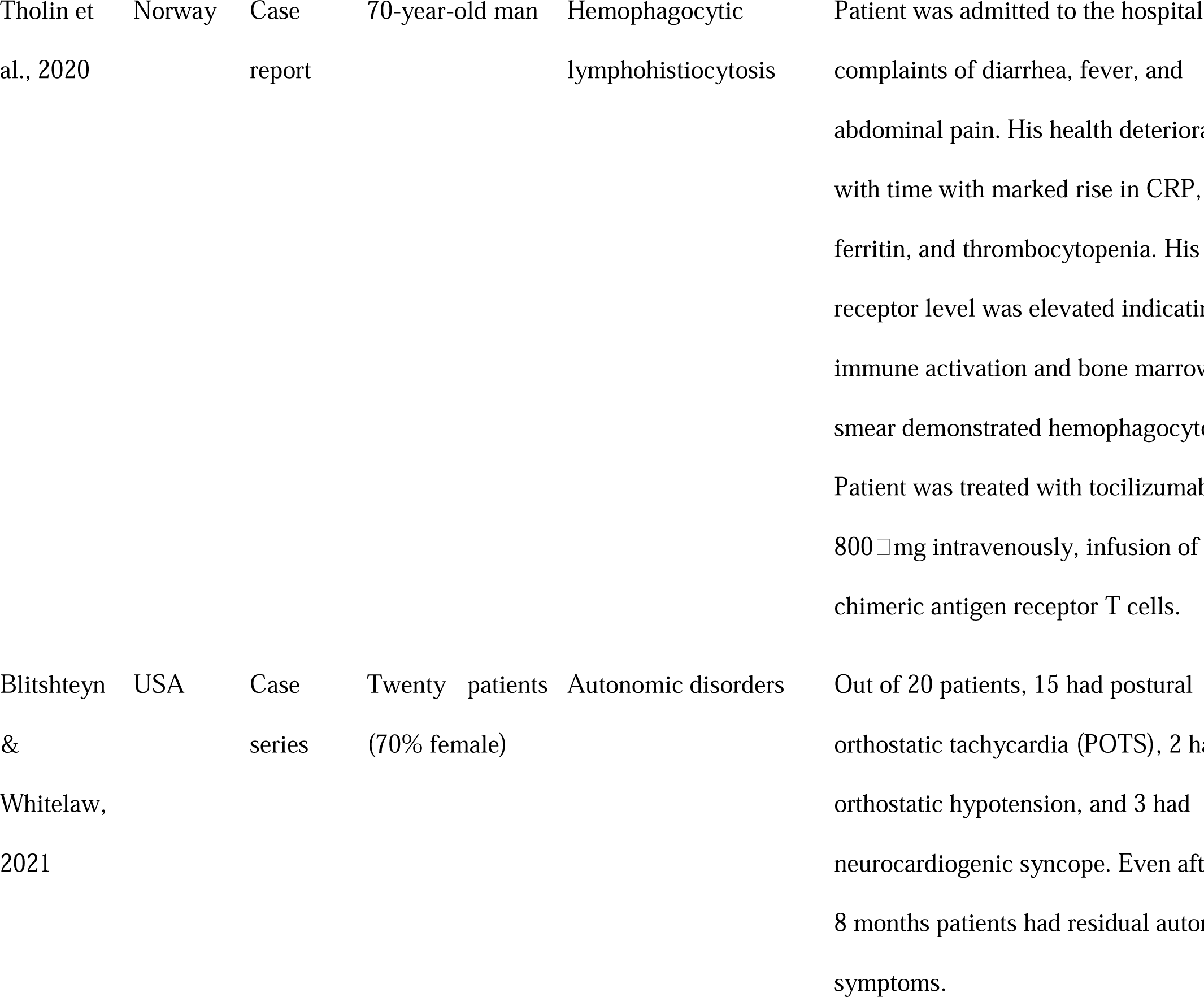
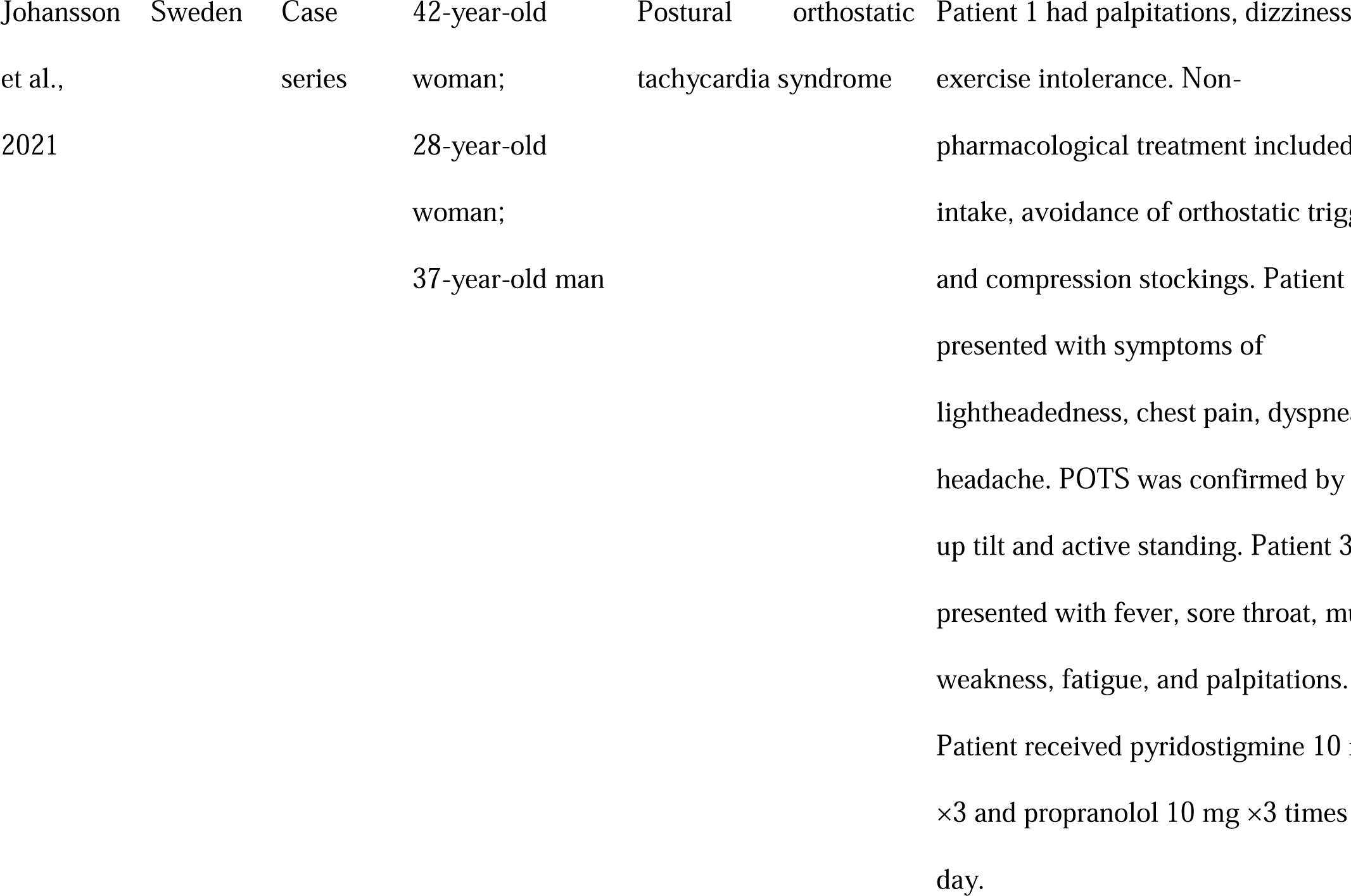
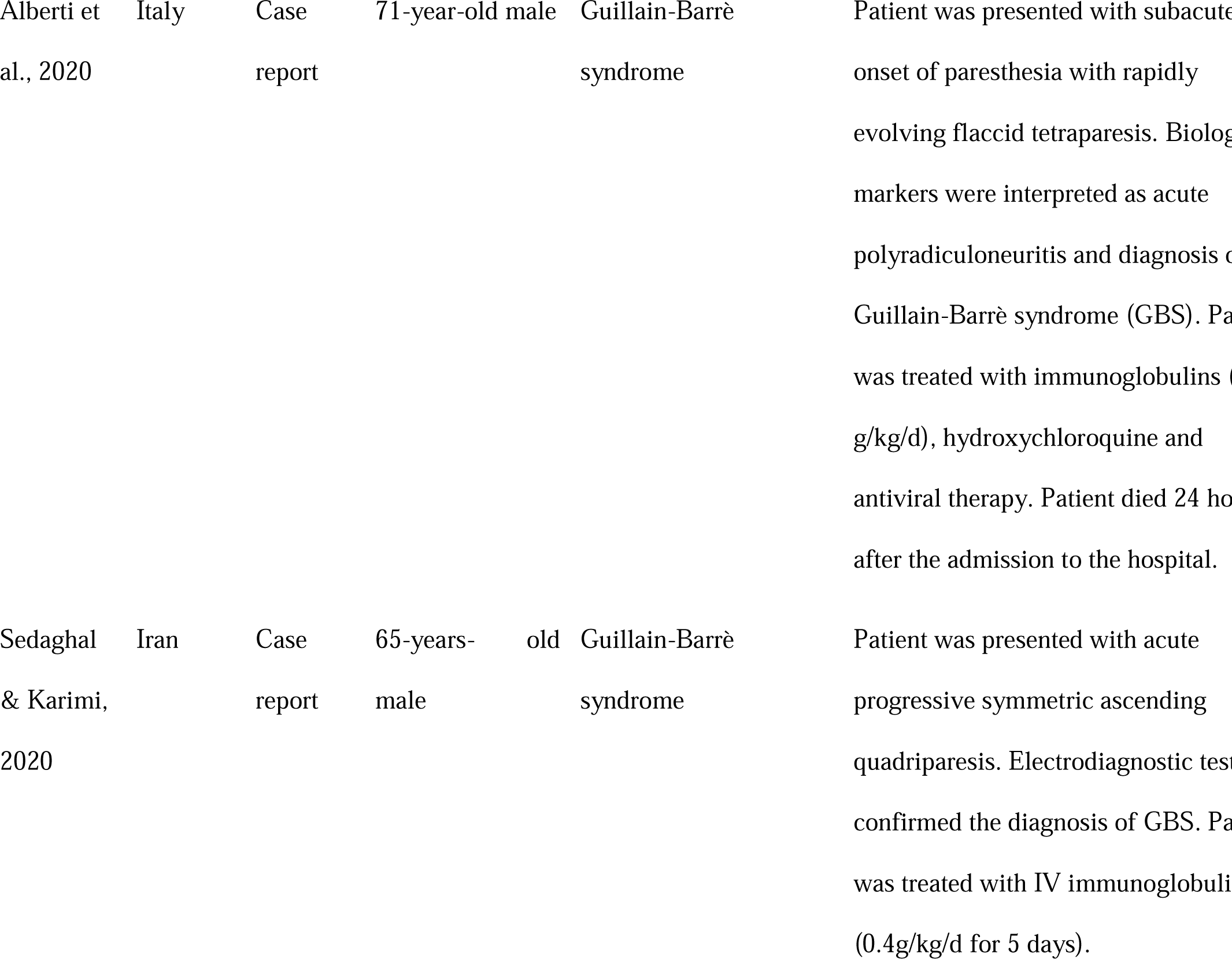
Data Extraction table.

## Results

### Characteristics of the study and sample

Twelve studies (three case series, eight case reports, and one retrospective cohort study) based on the inclusion criteria were included in the review. The studies that met the inclusion criteria were from different regions around the globe. Six studies were from European regions: France (Revuz et al., 2020), Turkey (Otluoglu et al., 2020), Norway (Tholin et al., 2020), Sweden (Johansson et al., 2021), and two from Italy (Nicolotti et al., 2021; Alberti et al., 2020). Two studies were from North America (Bennett et al., 2020; Blitshteyn & Whitelaw, 2021), four from Asia: two from India (Dhingra et al., 2021; Kalita et al., 2021), and one each from Lebanon (Naous et al., 2021) and Iran (Sedaghal & Karimi, 2020). A total of 36 patients constitute the sample size of the review. The age of the patients ranged from 28 years to 76 years, however, one study reported the case of a 2-year-old child experiencing post-COVID-19 syndrome (Kalita et al., 2021).

### Syndromes

All the included studies reported syndromes experienced by the patients after COVID-19 infection. The syndromes reported in the literature include immune thrombocytopenic purpura (ITP), viral encephalomyelitis, thrombotic thrombocytopenic purpura (TTP), hemophagocytic lymphohistiocytosis, postural orthostatic tachycardia syndrome (POTS), and Guillain-Barrè syndrome (GBS). Two case reports were on ITP (Revuz et al., 2020; Bennett et al., 2020). ITP diagnosis was determined based on reduced platelet counts, an increased ferritin level, and thrombocytopenia, as indicated in bloodwork. In both case reports of ITP, intravenous immunoglobulin (IVIG; 1 g/kg) constituted the first line of treatment and patients responded well to the treatment. One of the included case studies was based on viral encephalomyelitis experienced by patients after COVID-19 infection. Diagnosis was based on hyperintense lesions, found on MRI, on the surface of the temporal lobe and posterior medial cortex. Two cases of TTP were reported where the diagnosis was established based on: anemia, thrombocytopenia, reduced activity of ADAMTS13 and increased activity of anti-ADAMTS13 antibodies. Plasma exchange therapy was performed for both patients. One patient was given a fresh plasma transfusion followed by a five-day course of methylprednisolone (Nicolotti et al., 2021) while the other patient received cyro-poor plasma and received two doses of IM Vincristine and one dose of IM Rituximab (Dhingra et al., 2021). Both patients recovered and were discharged. Three of the included case reports presented the case of hemophagocytic lymphohistiocytosis (Tholin et al., 2020; Kalita et al., 2021; Naous et al., 2021). The diagnosis was made based on bone marrow smears in all cases. The patients were treated with steroids, antibiotics, and fluids. One patient was also infused with chimeric antigen receptor T-cells (Tholin et al., 2020). One of the patients did not survive despite high-dose steroid therapy and antibiotics (Naous et al., 2021). Other syndromes identified in the literature included POTS (Blitshteyn & Whitelaw, 2021; Johansson et al., 2021) and GBS (Alberti et al., 2020; Sedaghal & Karimi, 2020). For POTS, the diagnosis was confirmed using a head-up tilt test and measuring heart rate during active standing. Acetylcholinesterase inhibitors and beta blockers were for the pharmacological treatment of POTS in addition to non-pharmacological treatments such as abdominal binders, waist-high compression stockings, and fluids. For GBS, the diagnosis was based on biological markers suggestive of polyradiculoneuritis. IVIG constituted the first line of treatment. The details of each of the syndromes are presented as follows.

### Immune Thrombocytopenic Purpura

ITP is a bleeding disorder that is distinguished by a reduced number of platelets in the blood, known as isolated thrombocytopenia, where the platelet count is less than 150k/µ/L. It is a rare disease with a prevalence of 20 individuals per 1 million adults, typically affecting adults over the age of 50 (Michel, 2009). Pregnant women and those of childbearing age are also at increased (Provan & Newland, 2015). The disease course is generally more favourable in children than in adults. Children tend to achieve full remission sooner than adults, who often have a more chronic disease picture. Nonetheless, adults can experience spontaneous remission, which typically occurs during the initial months of diagnosis. Mortality is higher among older adults and patients who do not respond to initial treatment.

### Pathophysiology

The pathophysiology of the disease is unclear. Many hypothesize that ITP results from the development of IgG autoantibody which targets structural glycoproteins IIb-IIIa situated on the platelet membrane (Stasi & Newland, 2011). This makes platelets prone to the process of phagocytosis by Kupffer cells and splenic macrophages in the liver. However, these autoantibodies have been identified in only 40-60% of patients with ITP (Nazy et al., 2018).

### Clinical Symptoms

The initial suspicion and severity of ITP can be determined by evaluating the patient’s skin and mucous membranes, as well as asking them about their tendency to bruise or bleed minor trauma. As in other primary haemostasis defects, mucocutaneous bleeding and deeper organ bleeding may occur. Clinical signs of ITP include purpura, petechiae, and ecchymosis primarily in the upper and lower limbs. Petechiae may also appear in mucosal membranes, such as the nasal septum, hard palate, or gums, leading to epistaxis and gum bleeds. Spontaneous widespread hematomas can cause the platelet count to drop below 10k µ/L. Although fatal complications are rare, ITP can be implicated in overt gastrointestinal bleeding or intracerebral haemorrhage (Bohn & Steurer, 2018).

### Treatment

Clinical observation typically begins when the platelet count drops to 30k/µL in the absence of active bleeding. Treatment is initiated when active bleeding occurs. Glucocorticoids are considered the first-line treatment and typically involve the administration of Prednisone at a dose of 1 mg/kg PO OD. If glucocorticoid therapy fails, Rituximab at a dose of 375 mg/m2 IV once/week for a month is considered the second line of treatment. In cases of refractory ITP, agents such as Romiplostim (1–10 mcg/kg once/week) and Eltrombopag (25–75 mg once/day) are often used (Bohn & Steurer, 2018). While full remission can be achieved in approximately two-thirds of patients by splenectomy, this treatment comes with the risk of encapsulated bacterial infection and thrombosis. If patients do not respond to glucocorticoid treatment or experience severe bleeding, anti-D immunoglobulin (IG) or IVIG may be recommended.

### Viral Encephalitis

Encephalitis is a condition characterized by inflammation of the brain parenchyma, which can result in neurological dysfunction caused primarily by infection or autoimmunity. Diagnosis of encephalitis is typically made through the identification of inflammation in brain tissue specimens, but since inflammation in such specimens is not always directly indicated, indirect diagnoses must often be made through ancillary non-invasive tests, including cerebrospinal fluid analysis and neuroimaging. It is important to note that many other neurological conditions can lead to encephalopathy without causing any evidence of inflammation in the parenchyma. Encephalitis is generally suspected when symptoms of neurological dysfunction are observed, such as behavioral changes, focal deficits, decreased level of consciousness, papilledema, seizures, and headaches, alongside systemic manifestations such as rash, myalgia, arthralgia, lymphadenopathy, gastrointestinal symptoms, respiratory symptoms, or a history of exposure to risk factors such as animal bites, endemic areas, or exposure to ticks or insects (Costa & Sato, 2020).

### Pathophysiology

It has been suggested that a wide range of organisms can cause encephalitis, including protozoa, spirochetes, viruses, bacteria, fungi, and Rickettsiae. Among these, viruses are the most common cause of encephalitis worldwide; the most commonly implicated of which are cytomegalovirus (CMV), Herpes simplex virus (HSV1 and HSV2), varicella-zoster virus, chikungunya virus, Nipah virus, dengue virus, and enteroviruses (EVs). The pathophysiology of encephalitis can vary depending on the causative agent involved (Jain, Patel & Bhatt, 2014).

### Clinical Signs

The major clinical signs of encephalitis include altered mental status, characterized by lethargy, decreased, or altered level of consciousness, or personality changes lasting for more than a day with no other alternative cause found; fever greater than 38°C within 72 hours before or after presentation; partial or generalized seizures that are not completely attributable to the patient’s previous seizure disorder; recent onset of focal neurologic outcomes; neuroimaging suggestive of brain parenchyma abnormality; abnormalities detected in electroencephalography that are consistent with encephalitis and not attributable to any other cause.

### Treatment

The first line of treatment for suspected encephalitis includes correction and supportive treatment for autonomic dysregulation, hepatic and renal dysfunction, and electrolyte disturbances. It is also important to treat non-convulsive status epilepticus and seizures. If the diagnosis is not confirmed within 6 hours of admission, then it is recommended to start treatment with acyclovir at 500 mg/m^2^ TD for children and adolescents and 10 mg/kg TD for adults. Doses should be adjusted if the patient has a previous history of renal impairment (Costa & Sato, 2020).

### Thrombotic Thrombocytopenic Purpura (TTP)

TTP is classified as a type of thrombotic microangiopathy (TMA), a diverse set of disorders characterized by microangiopathic hemolytic anemia, thrombocytopenia, and organ dysfunction resulting from disturbed microcirculation. TTP is a rare disorder occurring in approximately 5 individuals per 1 million annually.

### Pathophysiology

In current literature, the pathophysiology of TTP is described as a severe deficiency of ADAMTS13, which can be the result of autoantibodies affecting its function, genetic abnormalities (congenital TTP), or dysregulated clearance of ADAMTS13 (autoimmune TTP). Persistent UL-VWF MM (ultra-large VWF multimers) results in the deficiency of ADAMTS13. UL-VWF MM with enhanced platelet aggregation occurs in the presence of stress-causing agents including, but not limited to, infections, pregnancy, surgery, and certain drugs. The aggregation of platelets causes an impedance in the flow of blood in the microcirculation, leading to clinical symptoms and organ damage. Although the central nervous system (CNS) is mainly affected by TTP, it also has an impact on other organs, including the heart and kidneys. In patients with acute TTP, platelets rich in von Willebrand factor with no or low fibrin have been identified in capillaries, small vessels, and large vessels (Lämmle, Hovinga & Alberio, 2005).

### Clinical Signs

Clinical signs of thrombotic thrombocytopenic purpura (TTP) and other thrombotic microangiopathies (TMAs) are characterized by disturbances in the microcirculation, consumption thrombocytopenia, and symptoms of red cell fragmentation (such as Coombs-negative hemolysis). Red cell fragmentation can be detected by free serum hemoglobin, elevated LDH, anemia, schistocyte count, reticulocyte count, hemoglobinuria, and reduced haptoglobin levels (Knöbl, 2013). Neurological symptoms resulting from brain hypoperfusion, such as blurry speech, headache, dizziness, amaurosis, epileptic seizures, stroke, or coma, can also occur due to TTP. Kidney involvement can lead to increased oligo- or anuria, serum creatinine, and hemolysis-induced hemoglobinuria. Thrombocytopenia may not always present with purpura and bleeding is rare, but hemolysis can cause anemia and jaundice. Cardiac involvement is a dangerous complication for patients with TTP (Mariotte et al., 2016).

### Treatment

Plasma exchange therapy is considered the first-line treatment for TTP and has been shown to improve the survival rate of 80-90% of patients. The therapy involves replacing 1.5 times the patient’s plasma volume with donor plasma. The donor plasma used can either be virus-inactivated plasma, fresh frozen single donor plasma, pooled donor plasma, or cryosupernatant. During therapy, the UL-VWF MM, autoantibodies, sludges, and immune complexes that are responsible for TTP are removed. This helps in reducing the severity of symptoms and improving the patient’s condition.

### Plasma infusion

It’s important to note that plasma infusion is not typically considered the first-line treatment for TTP. However, in patients with congenital TTP (a rare genetic form of the disorder), plasma infusion may be used as a treatment to replace the missing enzyme ADAMTS13. In these cases, regular prophylactic plasma infusions may also be necessary to prevent further episodes of the disease. The recommended dose for plasma infusion in TTP can vary depending on the severity of the disease and the patient’s individual needs.

### Immunosuppression

If TTP is autoimmune, then immunosuppression is achieved through corticosteroids such as prednisone 1-2 mg/kg to suppress the formation of other antibodies. In other forms of thrombotic microangiopathies, steroids are administered to reduce stress and enhance endothelial function (Zheng et al., 2020).

### Hemophagocytic Lymphohistiocytosis (HLH)

HLH is an uncommon but serious immunological syndrome characterized by elevated macrophage activity and cytotoxic lymphocytes, leading to multi-organ dysfunction and cytokine-mediated tissue injury. Several key soluble mediators including IL-18, interleukin (IL)-1b, and Interferon-gamma (IFN-γ) characterize HLH immunopathology. HLH has been classified into two forms: primary/familial form (F-HLH) and reactive/sporadic/secondary form. Familial HLH is conferred by the presence of penetrant genetic variation and mutation which affects lymphocyte survival, cytolytic function, and inflammasome activation. On the other hand, acquired factors such as infection, malignancy, and chronic inflammation constitute the basis for reactive HLH (Grom, Horne & De Benedetti, 2016).

### Pathophysiology

The underlying mechanism of HLH is yet to be elucidated. HLH is a distinct state of the activated immune system which can be achieved through several pathways depending on the environmental triggers and predispositions of the individual. Note that HLH is primarily driven by the abnormal immune system of an individual as opposed to underlying triggering agents. Moreover, the immune response in HLH, mediated by the activation of cytotoxic T-cells (CD8þ T-cells in particular) differs from the immune response in autoimmune disease as it does not target self-antigen agents. Chronic inflammation and immunosuppression also serve as predisposing factors for HLH. In such conditions, HLH is mostly triggered by viruses, but intracellular pathogens can also serve as triggering agents (Allen & McClain, 2015). Genetic mutations leading to HLH are clustered around the genes responsible for proteins implicated in lymphocyte activation and survival as well as cell-mediated cytotoxicity.

### Clinical Signs

Early diagnosis of HLH is crucial as patients can rapidly progress to multiorgan failure and death. Patients with HLH typically present with a persistently high fever, as well as other symptoms such as lymphadenopathy, hepatomegaly, splenomegaly, dysfunction of the CNS (including seizures or altered mental status), coagulopathy, and liver dysfunction. In some cases, patients may develop shock (Ramos-Casals et al., 2014). A common feature of HLH is a persistent high fever that does not respond to standard fever-reducing medications. Hepatomegaly is present in both children and adults with HLH, while splenomegaly is common but not present in all cases.

### Treatment

The treatment approach for HLH depends on the severity of the disease and the underlying triggering agent, if identified. The primary goal of treatment is to control inflammation, which is the underlying mechanism driving the disease. A multidisciplinary team approach is often necessary to provide comprehensive care for patients with HLH. Supportive care with blood products may be necessary for patients with abnormal blood clotting, and ventilator support may be required for critically ill patients with respiratory failure (Janka & Lehmberg, 2013).

### Postural Orthostatic Tachycardia Syndrome (POTS)

POTS is a clinical syndrome that presents with a variety of symptoms experienced by patients while standing, including palpitations, light-headedness, generalized weakness, tremors, exercise intolerance, blurred vision, and fatigue. A hallmark of POTS is an increase in heart rate of at least 30 bpm (or at least 40 bpm in individuals aged 12-19 years) within 10 minutes of moving from a reclining to a standing position. POTS is diagnosed when orthostatic hypotension, defined as a drop in systolic blood pressure of at least 20 mmHg, is absent. Presyncope symptoms are common in patients with POTS; standing heart rate is typically ≥120 bpm and higher in the morning than in the evening.

### Pathophysiology

Assuming an upright posture typically results in blood shifting from the chest to the legs and lower abdomen, and a significant volume of plasma moving from the vasculature to the interstitial space. This plasma shift can reduce venous return and lead to decreased stroke volume, cardiac filling, and arterial pressure. The compensatory sympathetic response, activated by baroreceptor signalling, can increase heart rate and systemic vasoconstriction, which restores cardiac output and venous return. In POTS, however, these physiological regulations are often compromised, resulting in reduced cardiac output, lack of normalization of cardiac volume, reduced venous return, and enhanced heart rate while standing. The underlying pathophysiology of POTS is not fully understood, but multiple factors likely contribute to the disorder, resulting in varied symptoms among individuals. Symptoms of POTS may result from excessive orthostatic shift in plasma volume, hypovolemia, increased sympathetic tone, enhanced blood venous pooling or poor venous return, physical deconditioning, autonomic dysfunction, and immunological factors (Wells et al., 2017).

### Clinical Signs

Orthostatic symptoms are the most frequent clinical manifestations of POTS, and they can be divided into non-cardiac symptoms (such as light-headedness, headaches/migraines, brain fog, nausea, weakness, tunnel/blurred vision, fatigue, and tremulousness) and cardiac symptoms (such as chest discomfort, exercise intolerance, dyspnoea, and heart palpitations). Light-headedness and presyncope are the most commonly reported symptoms in POTS, although only 30% of patients faint. It is important to note, however, that symptoms of POTS can also be non-orthostatic and general, including problems with sleeping, fatigue, migraines, daytime sleepiness, and hypermobility of joints (Mar & Raj, 2020).

### Treatment

The treatment of POTS involves both pharmacological and non-pharmacological approaches. Non-pharmacological treatments, which should be the first line of treatment, include exercise, increased salt and fluid intake, muscle tensing, using compression garments, modifying diet, and discontinuing medications that worsen the symptoms. POTS patients should also avoid stressors such as alcohol consumption, extreme heat, and dehydration.

Pharmacological treatments for POTS target the underlying cause of the disorder and aim to reduce heart rate, improve peripheral vasoconstriction, and increase intravascular volume. Midodrine and fludrocortisone are the most commonly prescribed medications, but their side effects may not be well-tolerated by some patients.

### Guillain-Barrè syndrome (GBS)

GBS is a rare, but serious, neurological disorder characterized by flaccid, acute, and neuromuscular paralysis. It was first described over a century ago, and since then, numerous studies have been conducted to investigate its presentation, immune-mediated pathophysiology, prognostic models, and treatment outcomes (Yuki & Hartung, 2012). GBS is typically classified as an “acute inflammatory demyelinating polyradiculopathy” because it affects the spinal cord and peripheral nerves, causing inflammation and damage to the myelin sheath that surrounds and protects nerve fibers. This damage can result in muscle weakness, arflexia, and sensory deficits, among other symptoms.

### Pathophysiology

GBS is considered to be an autoimmune disorder in which the immune system mistakenly attacks the myelin sheath covering nerve fibers in the peripheral nervous system (PNS). This results in inflammation, demyelination, and, in severe cases, axonal degeneration. The attack on the myelin sheath and nerve fibers leads to a breakdown in communication between the nerves and muscles, resulting in the characteristic flaccid paralysis seen in GBS (Leonhard, Ziemann, & Spies, 2021). The exact cause of GBS is not fully understood, but it is believed to be triggered by a preceding infection or vaccination in some cases.

### Clinical Signs

GBS is characterized by distal and proximal weakness, which can be flaccid and profound when the patient is hospitalized. As the disease progresses, patients may require intubation due to respiratory muscle weakness. GBS may also present with hyporeflexia and areflexia. Sensory symptoms, which are length-dependent, accompany the areflexia and flaccid weakness. Facial diplegia can also develop as both of the facial cranial nerves may be involved (Fokke et al., 2014).

### Treatment

The first-line treatments for GBS are plasma exchange therapy and IVIG. Plasma exchange therapy removes humoral mediators, pathogenic antibodies, and complement proteins, which are often pathogenic agents for GBS (Chevret, Hughes & Annane, 2017). IVIG exerts its therapeutic effects through immune-modulating actions.

## Discussion

The objective of the systematic review was to identify and characterize post-COVID-19 infection syndromes. The review identified six syndromes described in the literature, and a qualitative synthesis was conducted to synthesize the literature around their diagnosis and treatment methods. Here, we hypothesise potential mechanisms of how COVID-19 infection may lead to each of these syndromes.

Several pathways have been proposed that may lead to thrombocytopenia after COVID-19 infection, including the interaction of platelets with the virus through pathogen recognition receptors, hemophagocytosis caused by cytokine storms, sepsis, immune complexes and autoantibodies against platelets, platelet activation in lung tissue leading to coagulopathy, and microthrombi formation and damaged lung tissue leading to reduced platelet counts from megakaryocytes (Xu, Zhou & Xu, 2020). However, the most common mechanism leading to ITP is thought to be molecular mimicry between platelet glycoproteins and viral components. A study by Zhang et al. (2009) showed that protein sequences are shared between glycoprotein IIIa found on platelets and hepatitis C core-envelope peptides, which can trigger the production of antibodies that can fragment platelets. However, no sequence homology has been identified between platelet components and SARS-CoV-2 infection, so the exact mechanism remains unclear.

Three major pathways describe the underlying mechanism of SARS-CoV-2 infection causing encephalitis: the blood circulation pathway, direct infection injury, and neuronal pathways. Given that we know the functional receptor for the SARS-CoV-2 virus to be the ACE-2 receptor, which is present on the capillary endothelium, glial cells, and neurons, the blood circulation pathway suggests that these cells are thus potential targets for the virus, which may then enter the CNS through said receptors (Zhao et al., 2020). Another mechanism is the direct entry of the virus through the cribriform plate in the brain. Since the SARS-CoV-2 virus usually replicates in the nasopharyngeal epithelium, it can cause damage to neuronal tissue in the same manner as it does in olfactory nerves (Corona, Rodríguez-Violante & Delgado-García, 2020). Dynein and kinesin proteins in nerves, such as the vagus nerve, may also be responsible for the retrograde and anterograde transportation of the virus in the brain, facilitating insult via neuronal pathways (Zhao et al., 2020).

Viruses are known to commonly trigger thrombotic microangiopathies, but the exact mechanism leading to TTP from COVID-19 infection is unknown. However, several pathways have been proposed, including a high inflammatory state associated with cytokine storms, direct endothelial injury, or mediation via enhanced procoagulant factors such as von Willebrand factor, fibrinogen, and factor VIII (Panigada et al., 2020). Pascreau et al. (2021) observed 70 COVID-19 patients and determined their antigen levels, plasma VWF activity, and ADAMTS 13 antigen levels. They found a marked increase in VWF levels, which was associated with a decrease in ADAMTS13 levels, a pathognomonic indicator for TTP.

It has been proposed in the literature that the SARS-CoV-2 virus can cause cytokine storm including HLH, as there are many similarities in the clinical presentation of COVID-19 and HLH (Mehta et al., 2020). Poor outcomes have been observed for patients with SARS-CoV and MERS, which are associated with enhanced levels of proinflammatory cytokines (e.g., IL-1β) in body tissues, specifically in the lower respiratory tract (He et al., 2006). Increased levels of IL-1β enhance the production of other proinflammatory cytokines, including IL-6 and TNF-α, which can result in cytokine storm (Nieto-Torres et al., 2014). Therefore, SARS-CoV-2 may trigger secondary HLH in some patients.

The proposed mechanism for POTS post-COVID-19 infection is the dysregulation of the renin-angiotensin-aldosterone system (RAAS) due to the interaction of the SARS-CoV-2 virus with ACE2 receptors. ACE2 receptors are found on the endothelial cells of various organs including the lungs, heart, and kidneys; and we know that the virus binds to these receptors to enter the cell. This interaction can result in the downregulation of ACE2 receptors, which leads to an increase in angiotensin II levels, resulting in vasoconstriction, inflammation, and oxidative stress. This can lead to endothelial dysfunction and microvascular damage, ultimately leading to the development of POTS (Kanjwal et al., 2021).

It has been proposed in the literature that SARS-CoV-2 causes GBS syndrome in some patients either directly by binding with ACE2 receptors on neuronal tissue and manifesting their neuroinvasive capacity or indirectly through an autoinflammatory mechanism. Molecular mimicry may also play a role in the development of GBS in some COVID-19 patients. Molecular mimicry occurs when the immune system mistakes a viral protein for a self-protein, leading to an autoimmune response against the body’s tissues. A study by Toscano et al. (2020) reported that five COVID-19 patients with GBS had antibodies against gangliosides, which are molecules found on nerve cell membranes that can be targeted by the immune system in GBS. The antibodies were found to cross-react with the SARS-CoV-2 spike protein, supporting a potential role of molecular mimicry as the underlying mechanism. However, more research is needed to fully understand the mechanisms underlying GBS in COVID-19.

## Conclusions

Since the emergence of Coronavirus infectious Disease 2019 (COVID-19) in late December 2019, several vaccines have been made which subsided the pandemic; however post-COVID-19 infection, multiple syndromes have been reported globally. The intention of writing this systemic review is to summerise all the reported cases and hypothesised pathophysiology that might have led to the manifestation of the syndrome. Total of 12 studies were included which met the eligibility criteria where the reported studies were: immune thrombocytopenic purpura (ITP), viral encephalomyelitis, hemophagocytic lymphohistiocytosis, thrombotic thrombocytopenic purpura (TTP), Guillain-Barrè syndrome (GBS) and postural orthostatic tachycardia syndrome (POTS). Furthermore, presenting symptoms and treatment for each respective syndrome were discussed with brief background on coronavirus and its variants to provide a foundation on which to examine the syndromes manifested after COVID-19 infection.

## Data Availability

All data produced in the present work are contained in the manuscript.

## Declarations

### Authors’ contributions

SA conceptualised the systematic review and prepared the initial draft of the manuscript. AZ supported writing of the draft and oversaw its critical review and editing. *Both SA and AZ contributed equally to this work as co-first-authors. SK, AT, and KT undertook the literature search and helped prepare the manuscript draft. All authors read and approved the final manuscript.

