## Supplementary Materials for "Coronavirus and Post-COVID-19 Syndrome: A Systematic Review"

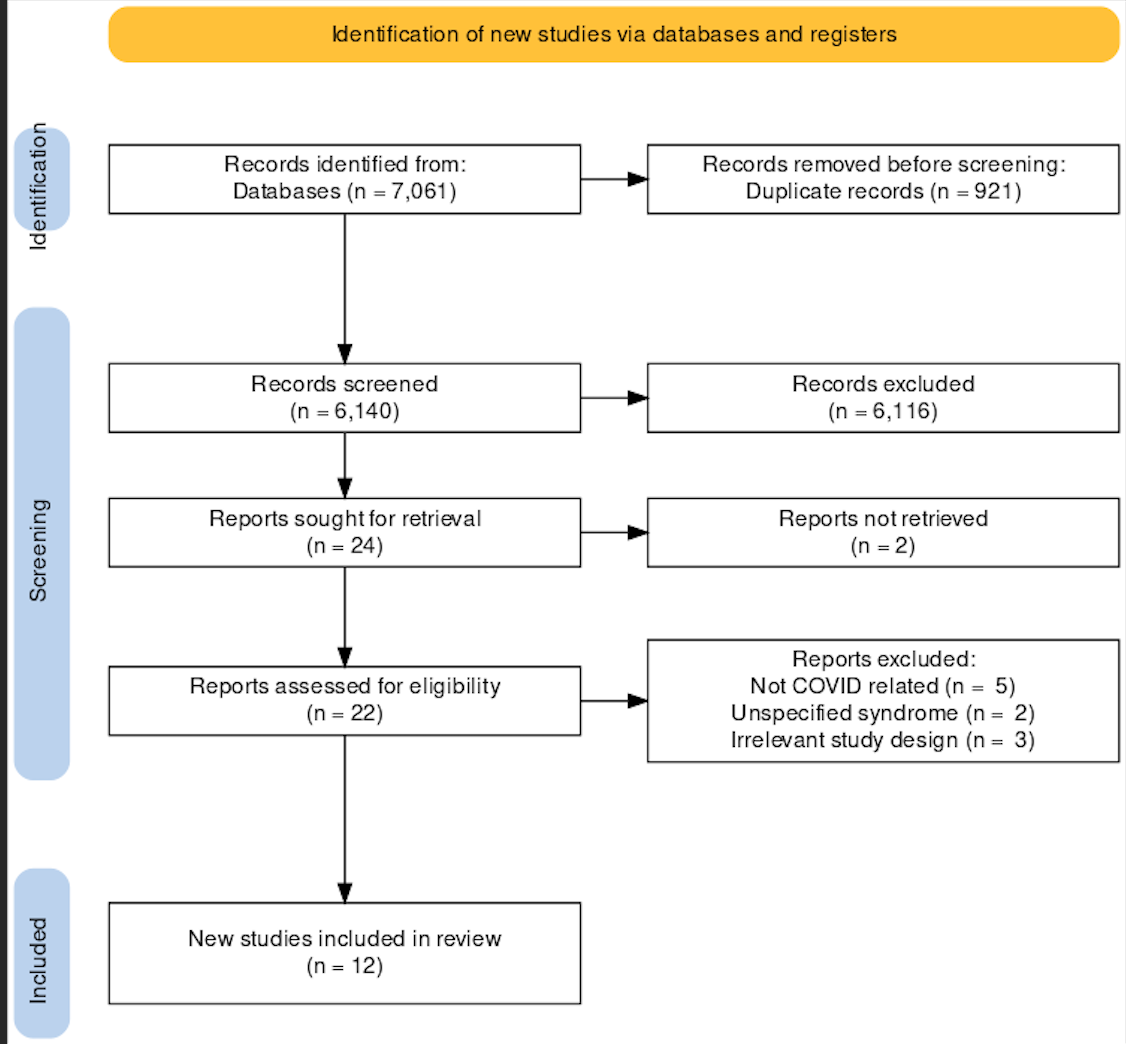

**Figure S1.** PRISMA flowchart outlining the paper screening process.

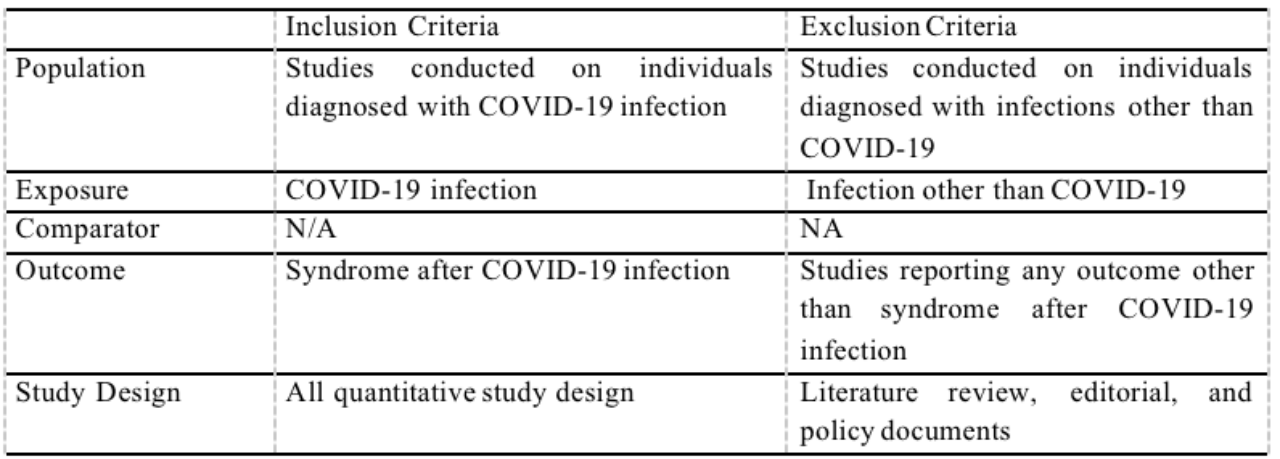
**Table S1.** Inclusion and Exclusion Criteria based on Population, Exposure, Comparator, and Outcome of Review Question.

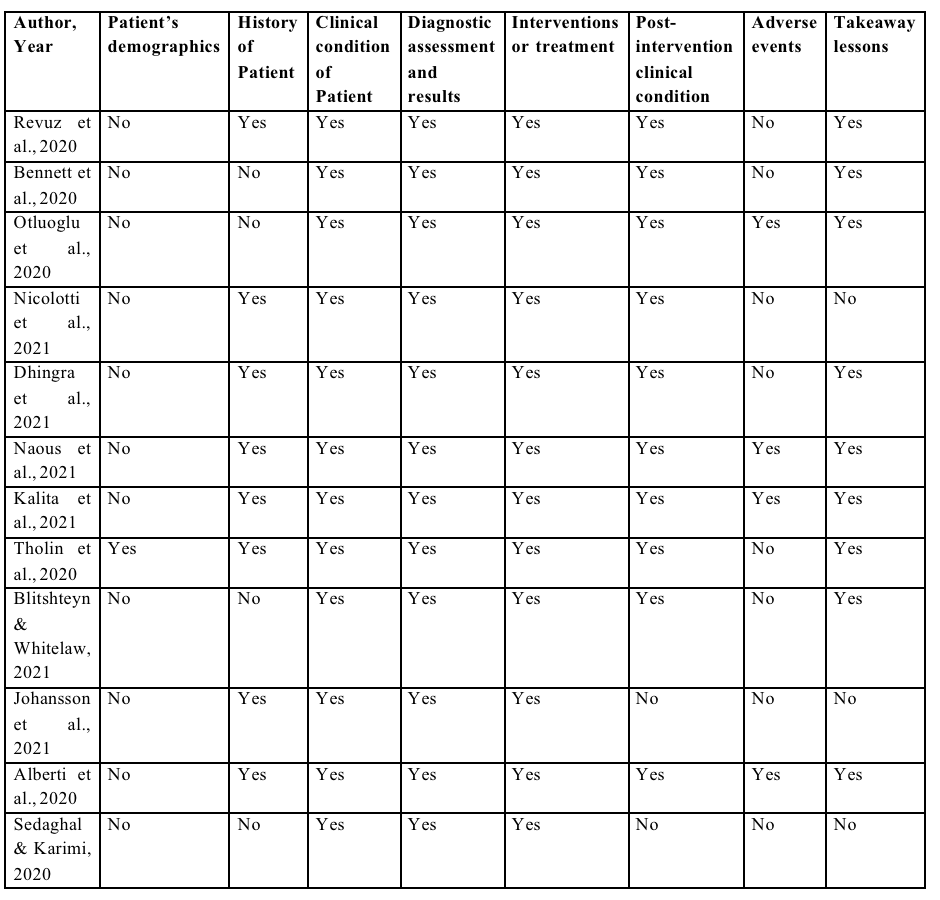
**Table S2.** Risk of Bias Assessment of included studies.
